# Artificial intelligence for diagnosis and prognosis in neuroimaging for dementia; a systematic review

**DOI:** 10.1101/2021.12.12.21267677

**Authors:** R Borchert, T Azevedo, A Badhwar, J Bernal, M Betts, R Bruffaerts, MC Burkhart, I Dewachter, HM Gellersen, A Low, L Machado, CR Madan, M Malpetti, J Mejia, S Michopoulou, C Muñoz-Neira, M Peres, V Phillips, S Ramanan, S Tamburin, H Tantiangco, L Thakur, A Tomassini, A Vipin, E Tang, D Newby, J Ranson, D.J. Llewellyn, M Veldsman, T Rittman

## Abstract

**Introduction:** Recent developments in artificial intelligence (AI) and neuroimaging offer new opportunities for improving diagnosis and prognosis of dementia. To synthesise the available literature, we performed a systematic review.

**Methods:** We systematically reviewed primary research publications up to January 2021, using AI for neuroimaging to predict diagnosis and/or prognosis in cognitive neurodegenerative diseases. After initial screening, data from each study was extracted, including: demographic information, AI methods, neuroimaging features, and results.

**Results:** We found 2709 reports, with 252 eligible papers remaining following screening. Most studies relied on the Alzheimer’s Disease Neuroimaging Initiative (ADNI) dataset (n=178) with no other individual dataset used more than 5 times. Algorithmic classifiers, such as support vector machine (SVM), were the most commonly used AI method (47%) followed by discriminative (32%) and generative (11%) classifiers. Structural MRI was used in 71% of studies with a wide range of accuracies for the diagnosis of neurodegenerative diseases and predicting prognosis. Lower accuracy was found in studies using a multi-class classifier or an external cohort as the validation group. There was improvement in accuracy when neuroimaging modalities were combined, e.g. PET and structural MRI. Only 17 papers studied non-Alzheimer’s disease dementias.

**Conclusion:** The use of AI with neuroimaging for diagnosis and prognosis in dementia is a rapidly emerging field. We make a number of recommendations addressing the definition of key clinical questions, heterogeneity of AI methods, and the availability of appropriate and representative data. We anticipate that addressing these issues will enable the field to move towards meaningful clinical translation.

## Introduction

There is a pressing need to improve the diagnosis and prognosis for people with dementia. Up to 20% of people may receive the wrong diagnosis (Fischer et al. 2017) and there is large geographic variability in the likelihood of receiving a diagnosis even within a single country (Cook, Souris, and Isaacs, 2019). Receiving a timely and accurate diagnosis is critical for people with dementia, their carers and families: it provides the opportunity for forward planning; and with the advent of disease modifying treatments an early accurate diagnosis will guide treatment selection, working towards precision medicine.

Neuroimaging is a non-invasive investigation used in routine clinical practice for the diagnosis of dementia in many countries (Rittman 2020; Filippi et al. 2012). A range of neuroimaging methods are used in dementia, of which computed tomography (CT), magnetic resonance imaging (MRI) and electroencephalogram (EEG) are widely available and relatively inexpensive methods to examine brain structure (Harper et al. 2014; Karas et al. 2003), longitudinal patterns of atrophy (Young et al. 2018) and changes in brain function (Pievani et al. 2011; Greicius et al. 2004; Stam et al. 2007; Badhwar et al. 2017). Positron Emission Tomography (PET) is available in specialist centres and is more expensive; it is used to measure metabolic activity or to identify specific underlying pathologies (Klunk et al. 2004; Lowe et al. 2016; Chételat et al. 2020). These neuroimaging methods have the potential to provide early clinical diagnostic and prognostic biomarkers for dementia.

Human clinical judgement and basic rating scales have traditionally been used to interpret clinical neuroimaging (Harper et al. 2014) using features such as medial temporal lobe atrophy (Scheltens et al. 1995) and white matter hyperintensity load (Wahlund et al. 2001; Fazekas et al. 1987). However, the development of more sophisticated approaches and richer data may mean that the most informative features are not amenable to human measurement.

Artificial intelligence (AI), and machine learning (ML) algorithms facilitate the automation of neuroimaging interpretation and have the potential to reduce bias and improve clinical decision making (Hainc et al. 2017; Kohoutová et al. 2020; Nielsen et al. 2020).

Neuroimaging data is particularly well-suited to analysis using AI given its high dimensionality, non-linear nature and high covariance within the data. Uncertainty remains about which AI/ML approaches have the greatest potential to inform clinical decision making and how their performance compares to human decision making.

A large and growing number of AI/ML studies have investigated how neuroimaging features can be used to predict cognitive diagnoses and conversion to dementia, fueled by the availability of large datasets, principally the Alzheimer’s Disease Neuroimaging Initiative (ADNI) (Mueller et al. 2005). Nonetheless, there are very few examples of AI/ML having a direct impact on clinical dementia services.

To make sense of this rapidly growing field and to identify the barriers to clinical translation, we systematically reviewed the current literature.

## Methods

We conducted a systematic review to investigate the use of AI and ML methods for diagnosis and/or prognosis in cognitive disorders secondary to neurodegenerative diseases including Alzheimer’s Disease (AD), Mild Cognitive Impairment (MCI), Parkinson’s Disease (PD), Vascular Dementia, Lewy Body Dementia (LBD), Frontotemporal Dementia (FTD), Progressive Supranuclear Palsy (PSP), Huntington’s Disease (HD) and Corticobasal Degeneration (CBD). A protocol for this review was registered with PROSPERO (ID: CRD42021232249) prior to the screening of abstracts. PROSPERO is an international database of prospectively registered systematic reviews where there is a health related outcome.

### Search strategy

The databases MEDLINE (via Ovid), Embase (via Ovid), Cochrane Library, BNI (via ProQuest), PsycINFO (via EBSCOhost), CINAHL (via EBSCOhost) and Emcare (via Ovid) were searched using the title, abstract, keyword and MeSH term fields from inception to January 8th 2021 with the support of the Cambridge University Library. Results were limited to English language studies. Full search terms for each database can be found in the supplementary material.

### Inclusion & Exclusion Criteria

The inclusion and exclusion criteria used during the screening process to determine which papers would be included in the systematic review can be found below:

Inclusion criteria:

● Primary research studies only.
● Patient population consisting of AD, MCI, PD, Vascular Dementia, LBD, FTD, PSP, CBD, HD and/or all-cause dementia.
● Involving at least one of the following neuroimaging or neurophysiological modality: structural or functional MRI, PET, single-photon computed tomography (SPECT), EEG, magnetoencephalography (MEG) or ultrasound.
● Used AI or ML methods to investigate diagnosis and/or prognosis of cognitive neurodegenerative disease(s).

Exclusion criteria:

● Studies which did not include human participants.
● Studies published in languages other than English.
● Conference abstracts, book chapters, articles which did not include primary research e.g. reviews.
● Studies where access to the full-text was not available despite attempts from multiple individuals involved in the screening process.
● Studies which did not use AI/ML methods or only used simple logistic or linear regression methods.
● Studies which combined neuroimaging with other biomarkers, such as cerebrospinal fluid (CSF) markers or genetics data, in the AI/ML algorithms without reporting of model performance for neuroimaging features without these other biomarkers.
● Studies which focused on automated segmentation techniques which did not directly relate to diagnosis/prognosis of neurodegenerative diseases.

### PICOS framework

Outline of the parameters of this systematic review according to the PICOS framework:

● <u>P</u>articipants: Patients with cognitive disorders due to neurodegenerative diseases.
● Index: Neuroimaging data assessed with AI and/or ML for diagnosis and/or prognosis.
● <u>C</u>omparator: Traditional manual/subjective diagnostic/prognostic assessment.
● <u>O</u>utcome: Accuracy of diagnosis and/or prognosis.
● <u>S</u>tudy design: Controlled study.

### Screening

A flow chart of the screening process according to PRISMA 2020 guidelines (Page et al. 2021), including the number of papers excluded at each stage, can be found in Figure 1. The initial records identified using the search criteria. These records underwent de-duplication using a Zotero (https://zotero.com) automation tool which flagged possible duplicate reports and were manually screened by a reviewer to merge genuine duplicates. Following de-duplication, all papers were screened across two stages. During the first stage, each abstract was independently reviewed by two reviewers to determine their eligibility for inclusion based on the outlined criteria using the screening tool Rayyan (https://www.rayyan.ai/). Once both reviewers screened their allocated abstracts, inclusion/exclusion decisions were unblinded. For abstracts where there was disagreement between screeners, a third independent reviewer assessed the abstract and made the final decision as to 1) progression to full-text screening stage or 2) exclusion.

**Figure 1:**
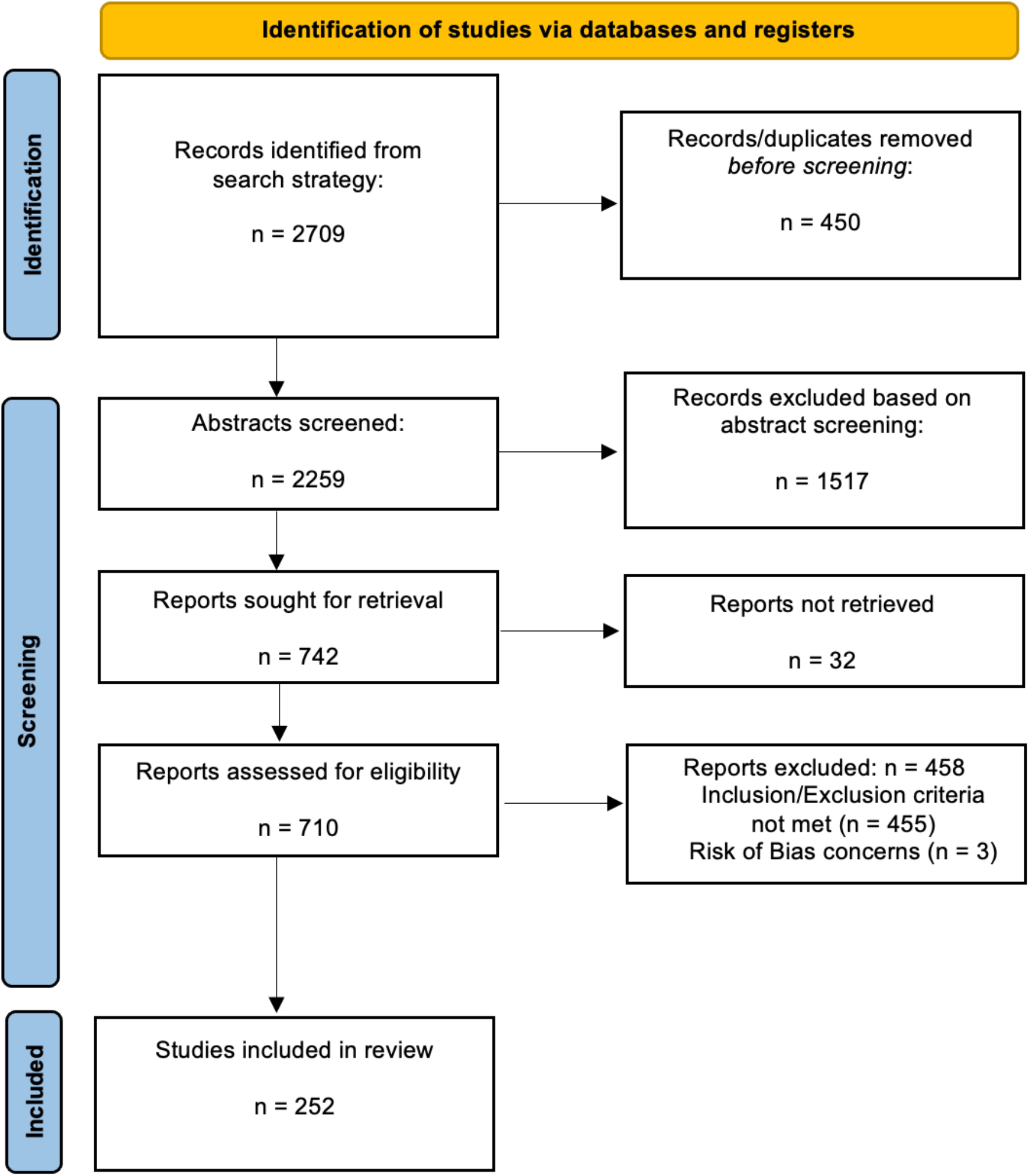
PRISMA 2020 flow diagram for systematic review outlining the number of reports identified and excluded at each stage.

The second stage involved full-text screening of all included papers by one reviewer per paper. For papers where the reviewer was unsure if the study met the outlined criteria, a second opinion was sought and a joint decision made after discussion with the second reviewer. Following the second stage of screening, all included papers were screened for risk of bias using the Joanna Briggs Institute (JBI) Critical Appraisal checklist (Moola et al. 2019).

### Data extraction

One reviewer per paper manually collected data from each report without the use of automation tools. The following data was extracted from the reports which passed the screening process:

1. Article information: First author, year, journal, country of first author’s affiliated institution.
2. Study method: Study design, patient population(s), control group(s), number of participants, neuroimaging modality, source of data. For reports using different datasets relating to a study, information regarding which specific dataset was extracted where possible. For example, for ADNI studies, the specific dataset used (ADNI-1, ADNI-2, ADNI-GO, J-ADNI) was identified and recorded where possible.
3. ML methods.
4. Receiver-Operator Curve (ROC) analysis results from the ML algorithm used to predict diagnosis/prognosis in the patient population including: accuracy (ACC), sensitivity (SEN), specificity (SPE), area under the curve (AUC), positive predictive value (PPV) and/or negative predictive value (NPV).

### Data analysis and approaches to classification

We used descriptive statistics to determine the following characteristics of the extracted dataset: source of neuroimaging data, type of neuroimaging used, AI/ML methods, focus on diagnosis and/or prognosis, accuracy of diagnostic/prognostic classifications, and global distribution of first authors’ institutions. The type of AI algorithm used for the diagnostic/prognostic classification task was extracted. Studies which used AI methods for feature extraction but not classification were excluded.

Given a training set of labelled features, there are multiple ways to learn a classifier that can then be used to predict class membership for new, unlabelled instances. We categorized classifiers according to the object they seek to learn or model.

1. Generative classifiers learn the joint distribution of the features and labels. Examples include Naïve Bayes and Linear/Quadratic Discriminant Analysis. After training, it is possible to generate (hence the name) new pairs of features and labels by sampling from the learned joint distribution.
2. Discriminative classifiers learn the conditional distribution of the labels given the features. Examples include k-nearest neighbors, and most ensemble methods (such as random forests).
3. Non-probabilistic, algorithmic classifiers directly learn the decision boundary in feature space. Examples include maximum margin classifiers and support vector machines.

We note that some non-probabilistic classifiers can be reframed in a probabilistic light (Franc, Zien, and Schölkopf 2011). For this reason, some authors consider these methods to be discriminative in nature and draw less of a distinction between our types (2) and (3).

In order to determine how well a classifier generalizes to new data, models are typically evaluated using a validation set consisting of labelled data withheld from the training process. The model’s predictions in the validation data can be compared to known labels using a variety of different metrics; precision, recall, accuracy, AUC, and F-scores are all estimated in this way. If a classifier performs much better on training data than on validation data, this can indicate overfitting. In such a case, the model may be refitted with regularization terms or priors that penalize model complexity.

Considering the large number of studies using ADNI and significant overlap of datasets, there is a risk of identifying spurious associations and false-positive findings when running a comprehensive meta-analysis (Pellegrini et al. 2018; LeBlanc et al. 2018; Han et al. 2016). We attempted to overcome these barriers by running a focused evaluation of the performance of AL algorithms, measured with AUC values, for a specific task: classification of AD vs healthy controls. This was achieved using a Stratified Weighted Sum (SWS) approach by assigning weights to the datasets and features (further methodological details in the supplementary materials).

## Results

The initial search strategy yielded 2,709 reports which were consolidated to 252 reports (Supplementary materials) after full-text screening (Figure 1). No additional items were included through analysis of the reference lists of the included papers. The publication time period of these papers ranged from 2005-2021.

### General characteristics of included reports

The final 252 included manuscripts were classified by country based on the institutional affiliation of the first author. The most common countries included China (25%), USA (17%), Italy (7%), France (6%) and South Korea (6%) (Table 1). Studies were also classified by their outcome measure(s), with the majority of reports focused on diagnosis of all-cause dementia (n=194) while a minority investigated both prognosis and diagnosis (n=22) or prognosis only (n=36).

**Table 1:** Number of included studies by country based on the institutional affiliation of the first author.

| Country of first author | Number of citations<br>(% total citations) |
| --- | --- |
| China | 64 (25%) |
| USA | 44 (17%) |
| Italy | 18 (7%) |
| France, South Korea | 16 (6%) |
| India | 10 (4%) |
| UK | 8 (3%) |
| Iran | 7 (3%) |
| Canada, Japan, Spain | 6 (2%) |
| Brazil, Germany | 5 (2%) |
| The Netherlands | 4 (2%) |
| Argentina, Colombia, Denmark, Egypt,<br>Finland, Switzerland, Turkey | 3 (1%) |
| Austria, Belgium, Portugal, Singapore | 2 (<1%) |
| Australia, Hungary, Jordan, Malaysia, Mexico,<br>Taiwan, Ukraine, Norway | 1 (<1%) |

### Datasets

Few studies used more than a single dataset, with 233 studies using one dataset, 16 used two datasets, and the remainder used three or more datasets. The most commonly used dataset was ADNI (see table 2). In the majority of the studies using the ADNI dataset the specific cohort used (ADNI-1, ADNI-2, ADNI-GO, J-ADNI) was not stated (see table 3). Where the cohort was available (n=52), 36 (69.2%) studies used a single cohort, 8 (15.4%) used two cohorts, and 8 (15.4%) used 3 cohorts. Of those that used ADNI-2 and ADNI-GO (n=11), a majority (n=9) also used ADNI-1.

**Table 2:** The majority of studies (70.6%) used the ADNI dataset alone or in combination with another dataset. Local data were used in 26.2% of studies. Multiple studies used a combination of two datasets or more resulting in an overlap between the categories listed here. ADNI = Alzheimer’s Disease Neuroimaging Initiative, OASIS = Open Access Series of Imaging Studies, AIBL = Australian Imaging, Biomarker & Lifestyle study of ageing, Bdx-3C = Bordeaux 3 Cities study, BLSA = Baltimore Longitudinal Study of Aging, CADDementia = Computer-Aided Diagnosis of Dementia challenge, NACC = National Alzheimer’s Coordinating Center.

| Dataset | N (%) studies |
| --- | --- |
| ADNI | 178 (70.6%) |
| OASIS | 5 (1.98%) |
| AIBL | 3 (1.2%) |
| Bdx-3C | 2 (0.8%) |
| BLSA | 2 (0.8%) |
| CADDementia | 1 (0.4%) |
| NACC | 1 (0.4%) |
| Others | 8 (3.2%) |
| Local data | 66 (26.2%) |

**Table 3:**
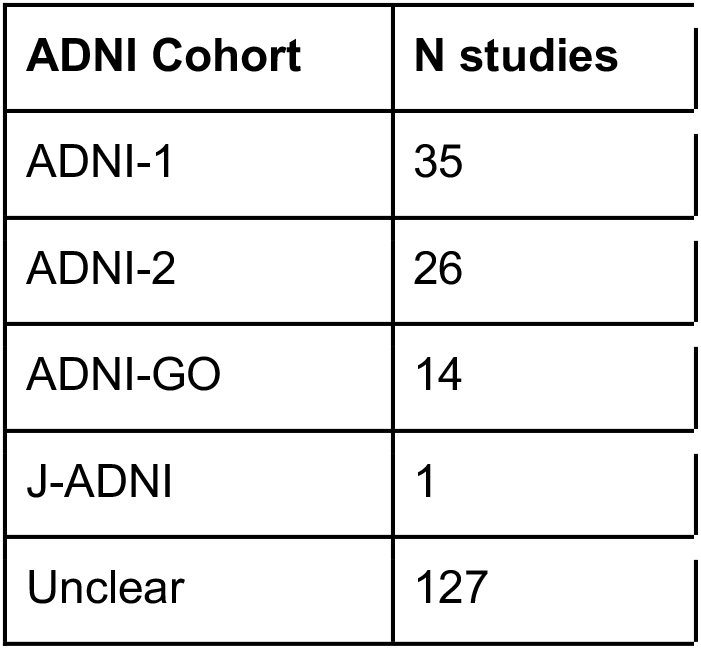
Specific cohort used in reports which included the ADNI dataset. ADNI = Alzheimer’s Disease Neuroimaging Initiative, J-ADNI = Japanese ADNI project.

Apart from using the ADNI dataset alone, 17 studies used data from ADNI combined with other datasets including the UK Biobank and AIBL. The majority (n=9) of these combination studies used a local dataset in addition to the ADNI dataset.

### Imaging modalities

The number of imaging modalities used across the included studies can be found in Figure 2. Structural MRI and PET/SPECT are the most frequent imaging modalities for diagnosis and prognosis of dementia, being used in approximately 71% and 26% of all works in the field, respectively. Around half of studies leveraged structural MRI alone (132/252) and those making use of multiple modalities (49/252) often used sMRI and PET (35/49) together. It is only since 2020 that studies incorporating three or more different modalities have begun to appear (T. R. Li et al. 2020; de Vos et al. 2020; Rabin et al. 2020).

**Figure 2.**
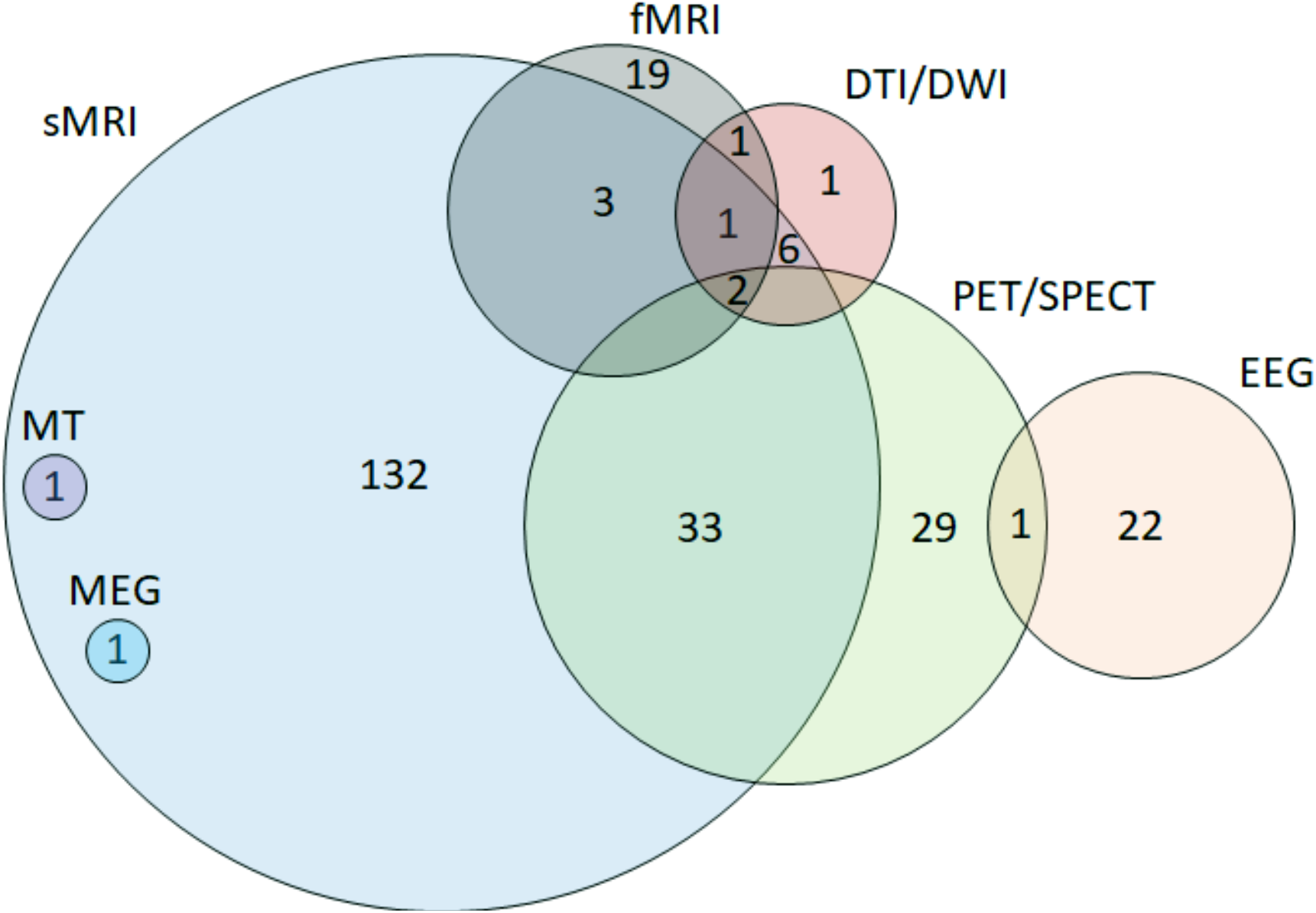
Imaging modalities used across included reports. fMRI = functional MRI, DTI = Diffusion Tensor Imaging, DWI = Diffusion Weighted Imaging, EEG = Electroencephalography, MT = Magnetisation Transfer, PET = Positron Emission Tomography, sMRI = structural MRI, SPECT = Single Photon Emission Computed Tomography, MEG = Magnetoencephalography.

### AI Methods

The classifier type most frequently used was a non-probabilistic algorithmic approach (47%), which mainly consisted of support vector machines (SVM), and discriminative classifiers (32%) which included most neural networks (Figure 3). Generative classifiers were used in 11% of studies while the “other” section (10%) mainly consisted of studies which combined multiple AI algorithms to generate novel, or complex classification tools which were difficult to categorise. From a qualitative perspective, most of these papers consisted of computationally focused studies which are not easily accessible to a clinical audience.

**Figure 3:**
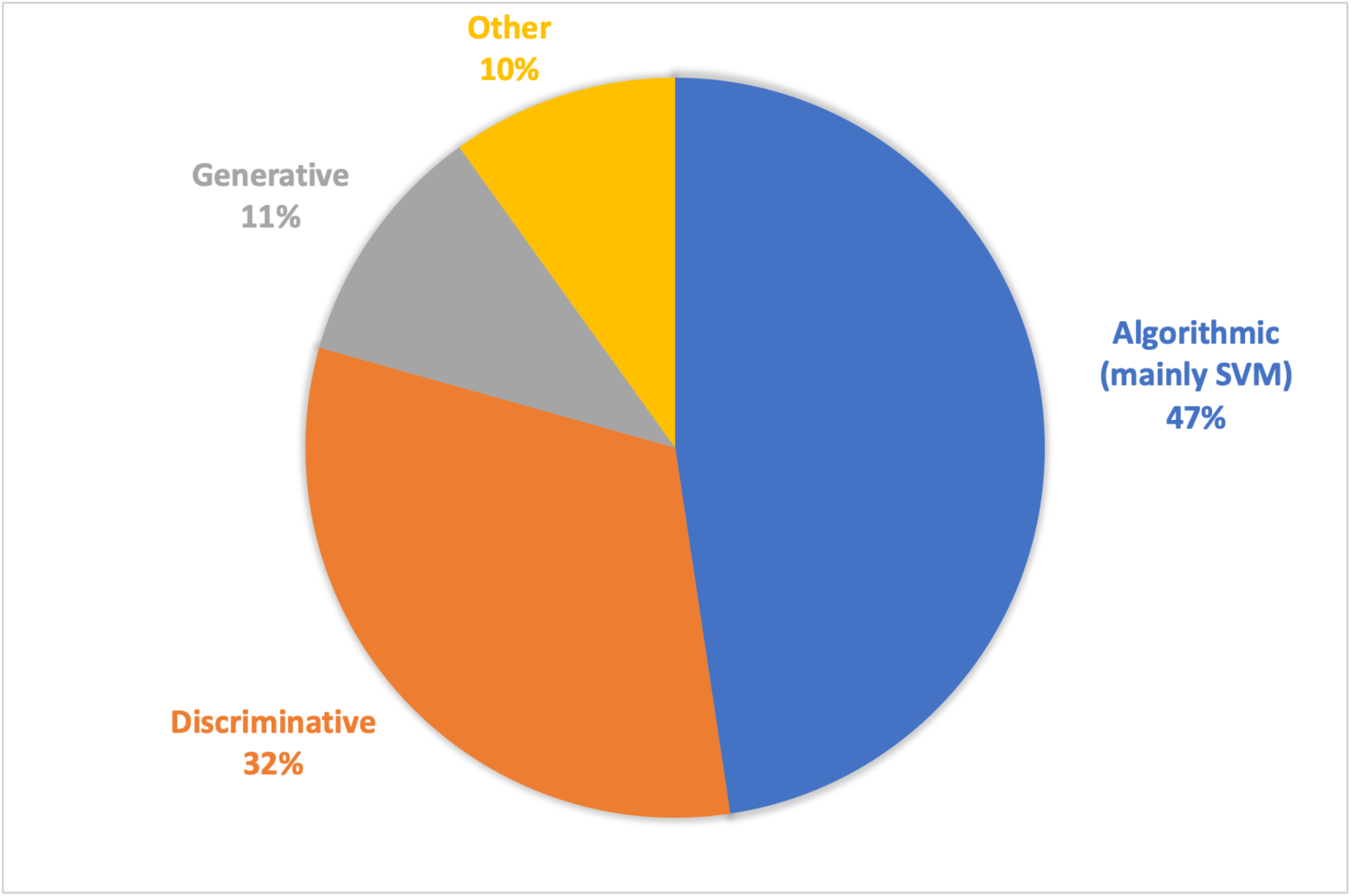
AI/ML methods used across the reports. Algorithmic classifiers (e.g. SVM) were by far the commonest method applied to neuroimaging data, followed by discriminative approaches (e.g. K-nearest neighbors, random forest). There was a wide range of other methods used including those which combined multiple classifiers.

The number of studies which used algorithmic classifiers (mainly SVM) increased considerably between 2013-2015 after which its use stabilised. In contrast, there was a sharp rise in the number of studies using discriminative approaches (mainly neural networks) to classification starting in 2017, with discriminative papers outnumbering algorithmic papers for the first time in 2019 (Figure 4).

**Figure 4:**
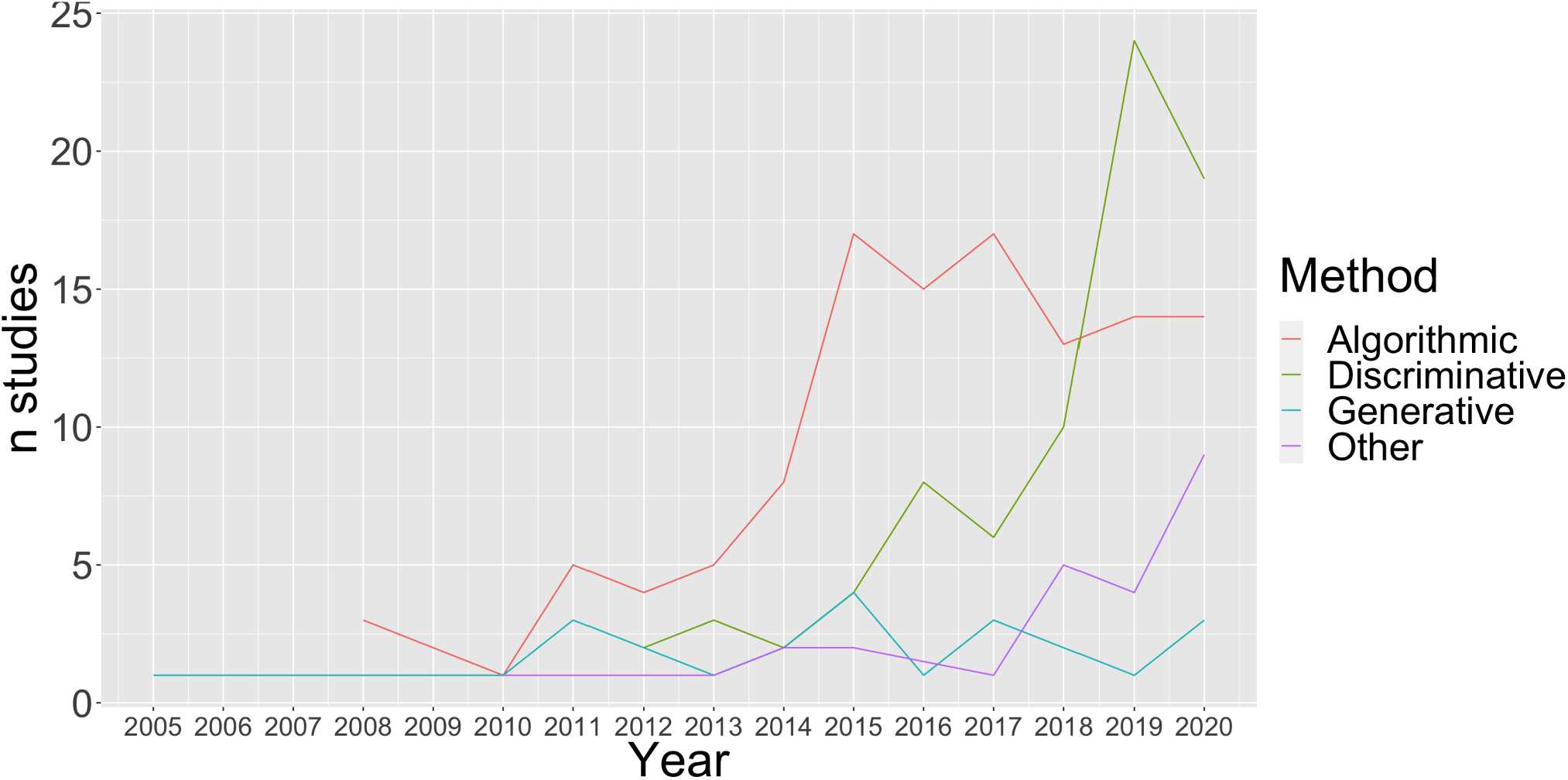
This figure shows the exponential rise in the use of discriminative classifiers, which mainly consist of use of neural network approaches, in the last 4 years. The use of algorithmic classifiers, predominantly SVMs, increased up to 2015 and has remained steady since. The use of generative models has stayed relatively stable since its first use in 2005 and other methods have been increasingly applied, particularly since 2017.

In order to unveil potential differences in performance between AI/ML methods, we examined AUC values for classifying AD vs healthy controls across studies (Figure 5). Note, only 13% (11/82) of these papers reported a confidence interval for the AUC value. Of these 11 papers, 5 did not report the level of confidence (eg 90% vs 95%).

**Figure 5:**
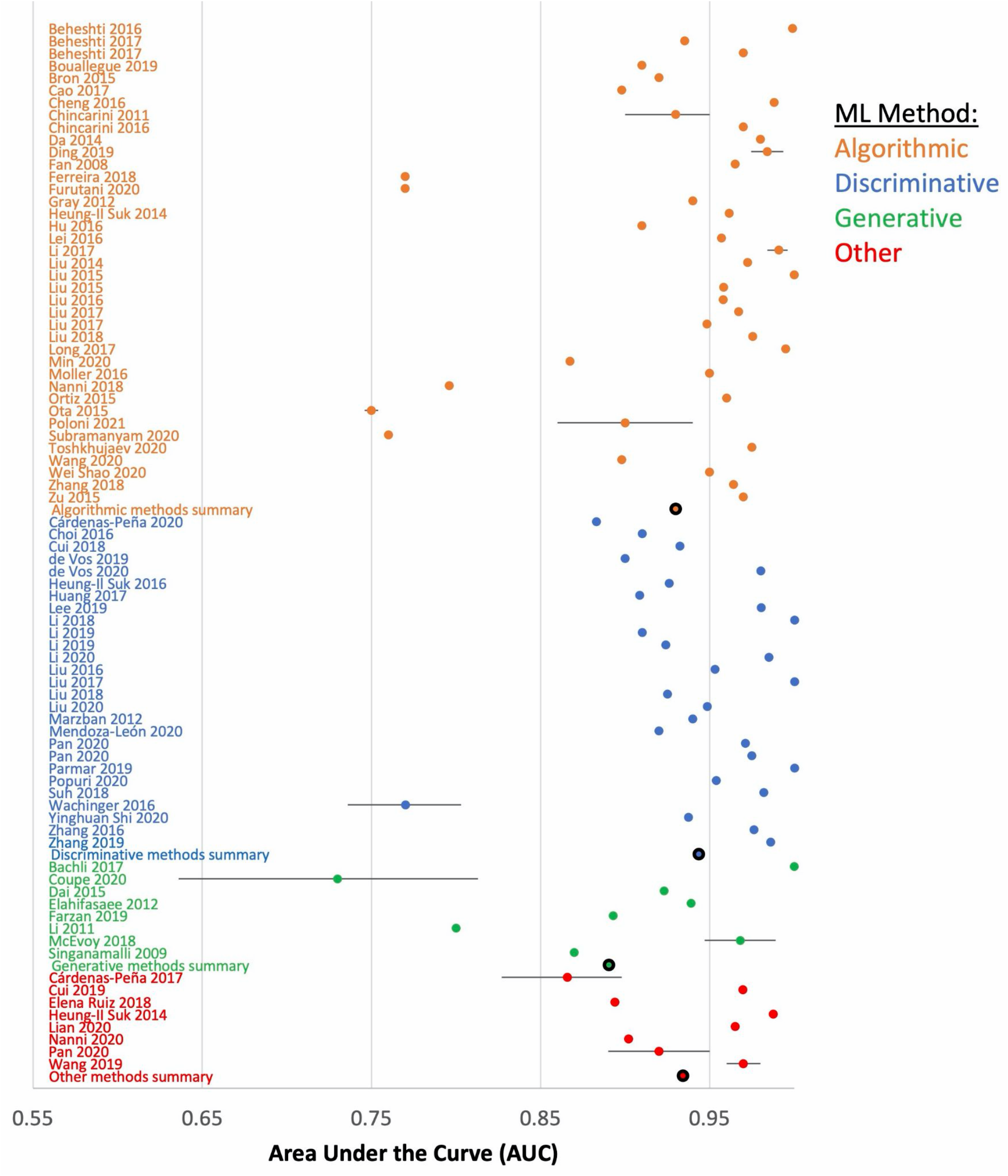
Forest plot depicting AUC values for classifications of AD patients versus healthy controls with confidence intervals where this was reported. Papers are stratified according to the type of ML method used including algorithmic (orange), discriminative (blue), generative (green) and other (red). Averaged AUC values for each type of ML method is depicted with a black border.

We employed a meta-analysis approach using the stratified weighted average sum (SWS) to weigh results based on the dataset, imaging modality, and type of AI/ML method used (methodological details in the supplementary materials). We found that for classification of AD vs healthy controls (i) discriminative models (SWS=0.602) performed better compared to algorithmic (SWS=0.585) and generative (SWS=0.556) classifiers; and (ii) the sparsity of the SWS matrix indicated that most of the literature was limited to only few datasets and imaging modalities.

We identified four studies which used transfer learning for classification (Jain et al. 2019; Lu et al. 2018; W. Li et al. 2020; Nanni et al. 2020). Transfer learning was typically used for fine tuning neural networks, particularly when the authors felt the dataset was not sufficiently large enough to properly train the neural network algorithm. Accuracy varied between these studies including for the following classification tasks: AD vs healthy controls (90.4 - 99.1), MCI vs healthy controls (83.2 - 99.2), and MCI converters vs non-converters (70.6 - 81.6).

### Diagnosis using volumetric structural MRI

In total 61% (153/252) of studies relied on volumetric structural MRI measurements. In the few studies that tested traditional and AI approaches head-to-head, AI methods outperformed raw volumetric measurements, for example using hippocampal volume for diagnosis (F. Li and Liu 2019; Morin et al. 2020) and predicting conversion of MCI to AD (Costafreda et al. 2011). The reported accuracy of AI methods for the diagnosis of Alzheimer’s disease varied between 60.2% to 99.3%. Of note, estimates in the lower range were found when using a multi-class classifier (i.e. AD vs MCI vs healthy controls, rather than AD vs healthy controls) (Guo et al. 2014; Cárdenas-Peña, Collazos-Huertas, and Castellanos-Dominguez 2017) or where an independent validation group was used (Kloppel et al. 2015).

Contributing to heterogeneity, the aim of “diagnosis” differed between studies using structural MRI. For example, there were 17 studies specifically targeting early diagnosis in which “early” disease was variably defined by: MMSE score < 24 (Cheng et al. 2017; Coupé et al. 2015; Gorji and Haddadnia 2015); CDR 0.5-1 (Dai et al. 2012; Hojjati, Ebrahimzadeh, and Babajani-Feremi 2019; Khedher et al. 2015; Nanni et al. 2020); progression from MCI to AD within 18 months (D. Pan et al. 2020; Salvatore et al. 2015), two years (Chincarini et al. 2011), three years (Hu et al. 2016; Moradi et al. 2015), conversion more than 12 months after imaging (Zhu et al. 2021); or was not clearly defined (H. Li and Fan 2019; Lisowska and Rekik 2019; Singanamalli, Wang, and Madabhushi 2017).

Studies using longitudinal structural MRI measures suggest that multiple timepoints may be more accurate than baseline measures alone for the diagnosis of AD (Farzan et al. 2011), and were particularly useful when applied to the prediction of MCI to AD conversion (Zhu et al. 2021; Cui, Liu, and Alzheimer’s Disease Neuroimaging 2019; D. Zhang, Shen, and Initiative 2012). Of interest, longitudinal changes in volumetric MRI may need to be considered in the context of baseline data to be meaningful (Chincarini et al. 2016).

### Other structural MRI methods

Twenty-eight studies investigated the use of non-volumetric structural imaging features for diagnosis (n=24) and/or prognosis/conversion (n=7). The input consisted of T1- or T2-weighted images, Diffusion Tensor Imaging (DTI) data or a combination thereof, to estimate non-volumetric features such as cortical thickness, texture and surface area. These studies focused on (i) optimization of image pre-processing techniques, (ii) investigation of feature selection methods, and (iii) optimization of classifiers and subsequent validation of the developed method. The accuracy for differentiating between AD patients and healthy controls ranged from 79.2% to 99.1%. Promising developments were noted for differential diagnosis (eg vascular dementia vs AD) (Castellazzi et al. 2020) and early diagnosis distinguishing MCI and healthy controls (Q. Li et al. 2018; Jung et al. 2015; Ebadi et al.

2017; Kruthika, Rajeswari, and Maheshappa 2019). Performance was lower when predicting MCI conversion to AD, or conversion of stable MCI to progressive MCI (Gao et al. 2020; Hett et al. 2018; Eskildsen et al. 2013).

### Functional MRI

Twenty-six studies (the first published in 2012) used resting-state MRI (rsMRI): we did not identify any studies using task-based MRI. All but four studies (Hojjati et al. 2017; 2018; Hojjati, Ebrahimzadeh, and Babajani-Feremi 2019; Rabin et al. 2020) focused on diagnosis and the majority (20/26) used ADNI data, either as the primary dataset or as a replication dataset. Graph measures were often used to summarize network characteristics. Overall, performance was satisfactory when discriminating between AD and controls (accuracies between 85-97%), but dropped when discriminating between MCI and controls (70-88%). Most studies reported the nodes which contribute most to discrimination between AD and controls: there is some heterogeneity, but most often components of the default mode network (DMN) were identified (Yang Li et al. 2019; Nguyen et al. 2019; Jin et al. 2020; M. Wang et al. 2020).

### Neurophysiological imaging

We identified twenty-four studies which used neurophysiological imaging methods, only three of which investigated non-AD dementias including PD and FTD (Garn et al. 2017; Ruffini et al. 2019; Dottori et al. 2017). The majority of the studies (n=21) used quantitative EEG while the remaining used either MEG (Furutani et al. 2020), event-related potential EEG (Chapman et al. 2011) or combined EEG with SPECT (Höller et al. 2017). Although half (n=12) of these studies were published since 2018, this cohort of publications also included some of the earliest papers identified in this review starting in 2005 (Dauwels et al. 2010; Cichocki et al. 2005). All neurophysiological studies used data from their local institution, the largest of which included EEG recordings from 272 participants (Buscema et al. 2010) although most studies (n=13) included less than 50 participants. Similarly to other imaging modalities, SVM was the most common (n=12) machine learning tool used and no other algorithm was used in more than three studies. Accuracy of discrimination between AD and healthy controls varied significantly from 69% in the single MEG study (Furutani et al. 2020) up to 100% in one study using four EEG features (Gallego-Jutgla et al. 2015).

### PET/SPECT imaging

Sixty-two studies were identified using PET imaging, aiming to improve early diagnosis (N=46), prognosis (N=10) or both (N=6) using AI/ML approaches. The most commonly used approach was SVM (N=22), which applied to FDG PET demonstrated an accuracy of >85% in studies for detecting AD hypometabolic patterns (Toussaint et al. 2012; Gray et al. 2012; De Carli et al. 2019) and outperformed structural MRI when compared head-to-head (Ferreira et al. 2018; Fan et al. 2008). Using SVM with FDG PET data distinguished AD (>86% accuracy) and MCI (>78.8% accuracy) from controls, and predicted MCI conversion within 12 months and up to 5 years with high accuracy (72%-80%) (Pardo et al. 2010; X. Pan et al. 2019; Ortiz et al. 2015; Y. Li et al. 2019; Teng et al. 2020; Cabral et al. 2015; Shen et al. 2019; Ota et al. 2015; Zhan et al. 2015; Ben Bouallegue et al. 2018; Yang et al. 2020; F. Liu et al. 2014). The same approach applied to amyloid PET also demonstrated accuracies of >85% for predicting MCI conversion and diagnosing AD (El-Gamal et al. 2018; Yang et al. 2020; Zhan et al. 2015; Xu et al. 2015; Nozadi and Kadoury 2018). Non-SVM approaches, such as convolutional neural networks and deep learning, on FDG PET and amyloid PET showed variable performance in predicting a final diagnosis of AD, cognitive decline or MCI conversion (Choi and Jin 2018; Ding et al. 2019; Huang et al. 2019; Son et al. 2020; Blazhenets et al. 2019; Morgado, Silveira, and Marques 2013; Popuri et al. 2018; Alzheimer’s Disease Neuroimaging Initiative et al. 2018; Lu et al. 2018; M. Liu, Cheng, and Yan 2018; Suk et al. 2015; F. Zhang et al. 2019), with accuracy between 75%-100%. Variability between studies could be explained by several factors, such as using whole-brain measures vs a priori selected regions, variable time intervals between follow-up visits, including vs excluding baseline cognitive scores in predictive models, and using data from multicentre studies (>70% accuracy) vs local databases (>78% accuracy).

Compared to ML methods which used PET alone, those which combined imaging modalities (i.e. FDG PET, amyloid PET and/or MRI) were more accurate in terms of diagnosis of both MCI and AD (Shao et al. 2020; Zu et al. 2016; Zhan et al. 2015; Ortiz et al. 2015; L. Liu et al. 2015; Yang et al. 2020; Xu et al. 2015; Ben Bouallegue et al. 2018). An additional approach used PET and structural MRI data in combination with other markers (i.e. APOE4 status and cognitive scores) to train a classifier, then selected neuroimaging features for classification, showing better performance when neuroimaging data (grey matter density, amyloid burden, APOE4 status; r = -0.68) were used to predict individualised rate of cognitive decline in MCI, compared to cognitive predictors (depression, memory and executive function scores; r = - 0.4) (Giorgio et al. 2020). Similarly, three studies showed that SPECT is able to classify MCI and AD, but its predictive value for MCI conversion is improved when combined with other imaging modalities or cognitive assessments (Borroni et al. 2006; Holler et al. 2017; Habert et al. 2011).

### Approaches to prognosis in Alzheimer’s disease

Fifty-six studies investigated either prognosis or a combination of diagnosis and prognosis. The majority were retrospective designs (49/52). Of 52 papers, 45 (87%) looked at prognosis in terms of MCI to AD conversion. Of these studies, two approaches were used to evaluate the performance of prognostic predictions; some exclusively used baseline data (fixed) while others used multiple imaging time points (continuous) and related these to time to conversion.

Thirty six papers only used baseline imaging data to predict a future diagnosis with a range of accuracy between 65% and 96% (mean AUC 0.79, standard deviation 0.09). Seven used multiple imaging time-points to make predictions with accuracies between 73% and 92% (mean AUC 0.81, standard deviation 0.10). One paper found a substantial improvement with longitudinal data (AUC 0.93) compared to baseline data alone (AUC 0.54) (Teng et al.

2020), and a second paper achieved a high level of accuracy using baseline neuroimaging information with longitudinal cognitive scores (AUC = 0.90) (H. Li and Fan 2019).

Time to conversion was divided into two categories: conversion within a fixed timeframe (40/45), or a continuous measure of time of conversion (5/45). Of those that used a fixed timeframe, 4 papers considered conversion within 1 year (AUC range: 0.73-0.90), 8 papers within 18 months (AUC range: 0.68-0.79), 5 papers within 2 years (AUC range: 0.74-0.96), 16 papers within 3 years (AUC range: 0.65-0.93), and 7 papers predicted conversion over 3 years with a maximum of within 10 years (AUC range: 0.54-0.91).

The main outcome of the remaining papers that did not focus on MCI to AD progression (7/52) varied; 2/7 predicted cognitive scores (Alzheimer’s Disease Assessment Scale - Cognitive Subscale/ADAS-Cog) over time using longitudinal MRI (Bhagwat et al. 2019; Zhou et al. 2013), while two other papers predicted both cognitive scores (Mini Mental State Examination/MMSE) and MCI to AD conversion within 24 months (D. Zhang, Shen, and Initiative 2012; Lei et al. 2019). Additionally, 2/7 papers predicted conversion from cognitively normal to AD in 7 (Coupe et al. 2015) and 2 years (Battineni et al. 2020). Finally, only one paper examined prognosis in a non-AD dementia, namely PD and DLB (Ruffini et al. 2019) with an AUC of 0.87.

MRI alone was the main imaging modality used (34/52 papers) with an additional six papers combining MRI and PET. Nine papers used only PET data, one used SPECT and two used EEG data. The main outcome measure for these papers was conversion to AD from MCI over a prespecified period of time (45/52 papers). A smaller proportion of papers (4 papers) used cognitive decline as an outcome measure. Similar to the diagnostic studies discussed in this review, the majority of the neuroimaging data came from the ADNI database (77%, 40/52 papers). An additional three papers combined local datasets with ADNI.

### Non-Alzheimer dementias

The majority of papers that included patients with non-Alzheimer’s dementia patients used neuroimaging features to improve the differential diagnosis between different dementia diagnoses. In total, 17 papers included a non-AD dementia group, fourteen featured a non-AD dementia as the diagnosis of interest, with the remaining three using the non-AD groups as a control group. FTD or behavioural variant FTD (bvFTD) was the most commonly investigated non-AD dementia, with seven papers having FTD or bvFTD as their main focus (Bachli et al. 2020; Cajanus et al. 2018; Dottori et al. 2017; Garn et al. 2017; Möller et al. 2016; Perry et al. 2017; J. Wang et al. 2016). These studies attempted differential diagnosis of FTD (from AD and/or LBD) most often using neuropsychological data and structural imaging (4/7 papers), with two papers using EEG (Dottori et al. 2017; Garn et al. 2017) and one using structural MRI for classification based post-mortem pathology (Perry et al. 2017). Five studies used data routinely collected in clinics (for example, from memory clinics) to attempt differential diagnosis between patient groups based on imaging features and typically included FTD, LBD, PSP, CBS, PDD and VAD (Bruun et al. 2018; Houmani et al. 2018; Kloppel et al. 2015; Koikkalainen et al. 2016; Morin et al. 2020).

Structural MRI was the most frequently used imaging modality (11/17 papers). Two papers focused on the differential diagnosis between PD and LBD (Oppedal et al. 2017; Ruffini et al. 2019) and only two on vascular dementia (Castellazzi et al. 2020; Ritter et al. 2016). The majority of papers used data from local hospitals or memory clinics (14/17 papers), one paper used local data combined with ADNI (Kloppel et al. 2015) and three papers used multi-centre or cohort data (Bachli et al. 2020; Bruun et al. 2018; J. Wang et al. 2016). Since the majority of studies utilised prospective or retrospective data from local clinics, datasets were relatively small compared to multi-centre studies like ADNI with most studies including 60-100 patients and some as low as 15 patients in a single diagnostic category (Castellazzi et al. 2020). The studies with larger patient numbers tended to come from multi-centre studies (Bruun et al. 2018; Bachli et al. 2020) or used retrospective data over a long period of time (Perry et al. 2017).

## Discussion

The growth of AI and ML promises a revolution in medicine, particularly in neuroimaging given its suitability for understanding large, complex data. We have reviewed the current state of the scientific literature using AI/ML methods applied to neuroimaging for clinical use in dementia. Key themes emerge, including a rapid rise in the use of neural networks and an increase in studies combining neuroimaging modalities. However, none of the algorithms reviewed are employed in clinics or registered as a medical device, highlighting that there remain obstacles to clinical use. The field is expanding rapidly with approximately 60% of the studies we included (n=157) published since the most recent similar review including studies up to 2016 (Pellegrini et al. 2018). Progress has been made in the last five years on some of the concerns raised by Pellegrini and colleagues, including the overreliance on SVM classifiers and MRI. However, despite this surge in studies, we are concerned that there are several barriers preventing the integration of these novel methods into clinical practice. Below we discuss three critical issues identified from this structured review: 1) defining key clinical questions, 2) the heterogeneity of methodological approaches, 3) the need for appropriate and representative data for validation.

### Defining key clinical questions

In order to be relevant to clinicians and patients, AI/ML algorithms must be used to solve questions that arise from the memory clinic. In broad terms, relevant clinical questions can be split into early diagnosis, differential diagnosis, prognosis and predicting response to treatment. There were no studies investigating response to treatment, unsurprisingly given that the currently widely available treatments for dementia are symptomatic rather than disease modifying.

The majority of studies considered the diagnosis of AD, and of those a large number considered prognostic prediction of MCI conversion to AD. However, variability in the diagnostic criteria applied limits the comparison between studies. For example, in studies which attempt to classify participants based on AD diagnosis, “early” disease was variably defined using neuropsychological testing, clinical acumen, conversion from MCI within an arbitrary time-frame, or was not defined at all. A single, clear definition of MCI and conversion to AD would be ideal, but has remained elusive despite recent efforts to reach such a consensus (Dunne et al. 2021).

It is encouraging to see increasing numbers of non-Alzheimer dementia studies using AI/ML for neuroimaging. The challenge for this group of diseases is the lack of a large, readily available dataset similar to ADNI for AD. Perhaps the most surprising is the lack of high quality data for vascular dementia, with only two studies included in the review and no dedicated dataset available.

We did not find any studies that assessed the common clinical challenge of differential diagnosis from among multiple (>2) possible diagnoses. This is a much harder problem to solve for AI/ML algorithms because it requires a multi-class classifier which is computationally more challenging and typically yields lower accuracy than a two-class classifier. Differential diagnosis also requires appropriate datasets. The ADNI dataset consists almost exclusively of MCI or AD patients, to the exclusion of other types of dementia. The NACC dataset in particular has larger numbers of non-Alzheimer dementia patients from a real world setting (Beekly et al. 2004), but is under-utilised, perhaps because of the variability in scanning sequences (including MRI field strengths), and the reliance on clinically defined diagnoses rather than research diagnostic criteria.

Another challenge to the clinical translation of AI/ML algorithms is the number of studies that are purely methodological, often without a clinician on the author list. These papers tend to be written with a mathematical focus, so are inaccessible for the majority of memory clinic professionals. This risks training an algorithm that answers the ‘wrong’ clinical question, and is therefore unlikely to progress any further. It is possible that the best performing algorithm exists within this set of studies, but may not be identified because it has not been developed as a clinical tool from the outset.

To ensure a clinical focus and clinically relevant hypotheses, we advocate the early and meaningful involvement of a clinician with expertise in dementia. To go a step further, we call for proper engagement of people with dementia and their relatives to identify questions in the field relevant to them. To address a broader range of dementia, we call for the establishment of large datasets of non-Alzheimer dementias, particularly including vascular dementia, and preferably including more than one type of dementia.

*Methodological heterogeneity*

The range of AI/ML algorithm types applied to neuroimaging and dementia represents a welcome growth of interest in the field. However, it introduces new challenges of making reliable comparisons between studies. This is exacerbated by the heterogeneity in diagnostic definitions and reporting.

There is some progress to address methodological heterogeneity within the broader neuroimaging field, such as the introduction of a reproducible classification protocol for MRI and PET data from the ADNI, ABL and OASIS datasets with the aim of improving reproducibility, accuracy and transparency (Samper-González et al. 2018). However, further efforts are needed to transition towards an agreement in the field on standard definitions, outcome measures and reporting to enable replication and meaningful comparison between studies.

For outcome in diagnostic classification ROC curve analysis is widely used, in particular we found the AUC is often reported as the main measure of accuracy, usually accompanied by the PPV and NPV. The PPV and NPV are more relevant to clinical practice, providing interpretation of the likelihood that the classification is correct given a positive or negative result. The outcome measure for prognostic studies is more challenging. We found that studies predicting prognosis usually grouped outcomes and applied ROC curve analysis. This is particularly relevant for predicting MCI to AD conversion, however it is not applicable to other situations, such as predicting the rate of cognitive decline in established dementia. Similar issues have been faced in other fields, most notably in cancer where the use of odds ratios, survival curves are encouraged, and pre-registering analysis (Riley et al. 2003). The idea of preregistering analysis is increasingly common and facilitated by structures such as the Open Science Framework (Sullivan, DeHaven, and Mellor 2019). It has been advocated for in both neuroimaging studies (Gentili et al. 2021) and AI/ML methodologies (Hildebrandt 2018).

The lack of reproducibility and transparency is an increasingly complex issue. Addressing these issues would be aided by availability of the source code. The limited descriptions of AI tools and code availability makes the prospect of reproducing most studies difficult at best, and in many cases impossible. Reproducibility is a challenge for many areas of science, including neuroimaging research, where statistical errors, small sample sizes and lack of detailed methods descriptions are common problems (Poldrack et al. 2017). The neuroimaging field has led the way in open science, such as data sharing and reproducibility through the Human Connectome Project (Van Essen et al. 2013). However, with the growing interest in commercializing AI tools for clinical practice, we speculate that transparency will become an increasing source of tension between academics promoting open science and private industry protecting intellectual property. Nevertheless, for the field in its current state of uncertainty, we believe that reproducibility within the scientific community trumps commercial interests to identify the best available AI/ML methods.

With the increased use of neural networks and model complexity comes reduced transparency and interpretability. Kohoutová and colleagues (Kohoutová et al. 2020) propose an staged approach to address some of these methodological issues in AI applied to neuroimaging by suggesting assessment of an algorithm at each of three levels: model level, feature level and the biological level. Such a staged assessment would provide a unified approach to compare between distinct algorithms with similar aims.

We call for future studies to report methodologies in full and provide sufficient information and source code to permit independent validation. We recommend that all studies of AI/ML in neuroimaging for dementia be pre-registered to ensure consistency and reliability of outcome measures. We strongly encourage the use of staged approaches to model validation.

### Data availability and representativeness

Closely linked to reproducibility is the question of generalisability, that is whether an AI/ML algorithm works as well in a completely independent dataset. This relies critically on the availability of appropriate data. External validation is hampered by the dominance of the ADNI dataset for training and testing. The widespread use of ADNI has undoubtedly accelerated the application of AI to MRI data in particular. Results from EEG studies are promising with classification comparable to MRI studies, yet have undoubtedly been hampered by the absence of an equivalent dataset.

The over-reliance on a single dataset such as ADNI from one country introduces potential ethnic and socio-economic biases to models that may hamper generalisation, an issue that has been specifically raised in the ADNI dataset (Mendelson et al. 2017). Concerns have been raised more generally about bias in AI/ML models (Sun, Nasraoui, and Shafto 2020), including in the context of health applications (Parikh, Teeple, and Navathe 2019). This is of particular concern in marginalised ethinic groups who have poorer health indicators in general (Williams 2012), and who may miss out on access to health services due to socio-demographic, cultural or religious beliefs (Obermeyer et al. 2019), including dementia services (Razai et al. 2021; Mukadam, Cooper, and Livingston 2011). More representative datasets are critical for models to translate reliably to all parts of the population, to inform risk prediction models and work towards closing gaps in health inequality related to dementia. A number of methodological approaches are available for measuring or mitigating bias (MehrabiNinareh et al. 2021), but we did not find the issue of bias to be discussed or addressed in the studies we reviewed.

We found that studies using an independent dataset for validation, as opposed to cross-validation or other similar methods, reported much lower accuracy, particularly when a community based population was used. For instance, applying an SVM classifier trained on ADNI and applied to memory clinics found markedly reduced accuracy in the clinical setting (AUC = 0.76 for AD diagnosis) compared to that in the training dataset (AUC=0.96) (Kloppel et al. 2015). A few recent studies have addressed the risk of overfitting by assessing generalisability in unseen independent research datasets (Sorensen and Nielsen 2018; De Carli et al. 2019; Qiu et al. 2020), collectively demonstrating the value of this approach in identifying methodological issues relevant to the overall model performance. Therefore, validation studies are critical, particularly those in a memory clinic setting where the tools are ultimately to be used.

In order to improve the availability of appropriate data, firstly we call for the establishment of a large, open EEG dataset for the development of AI/ML methods. Secondly, we call for each dataset to accurately represent the data from which it is drawn and that the community works towards datasets that represent the global population more accurately. Thirdly, we call for validation of AI/ML methods in real world memory clinic datasets.

### Next steps

To accelerate the translation of the innovative AI/ML approaches to diagnosis and prognosis in dementia identified in this review, we have devised a set of recommendations shown in Table 4 which are grouped into those for individual studies and challenges for the field in general.

**Table 4:**
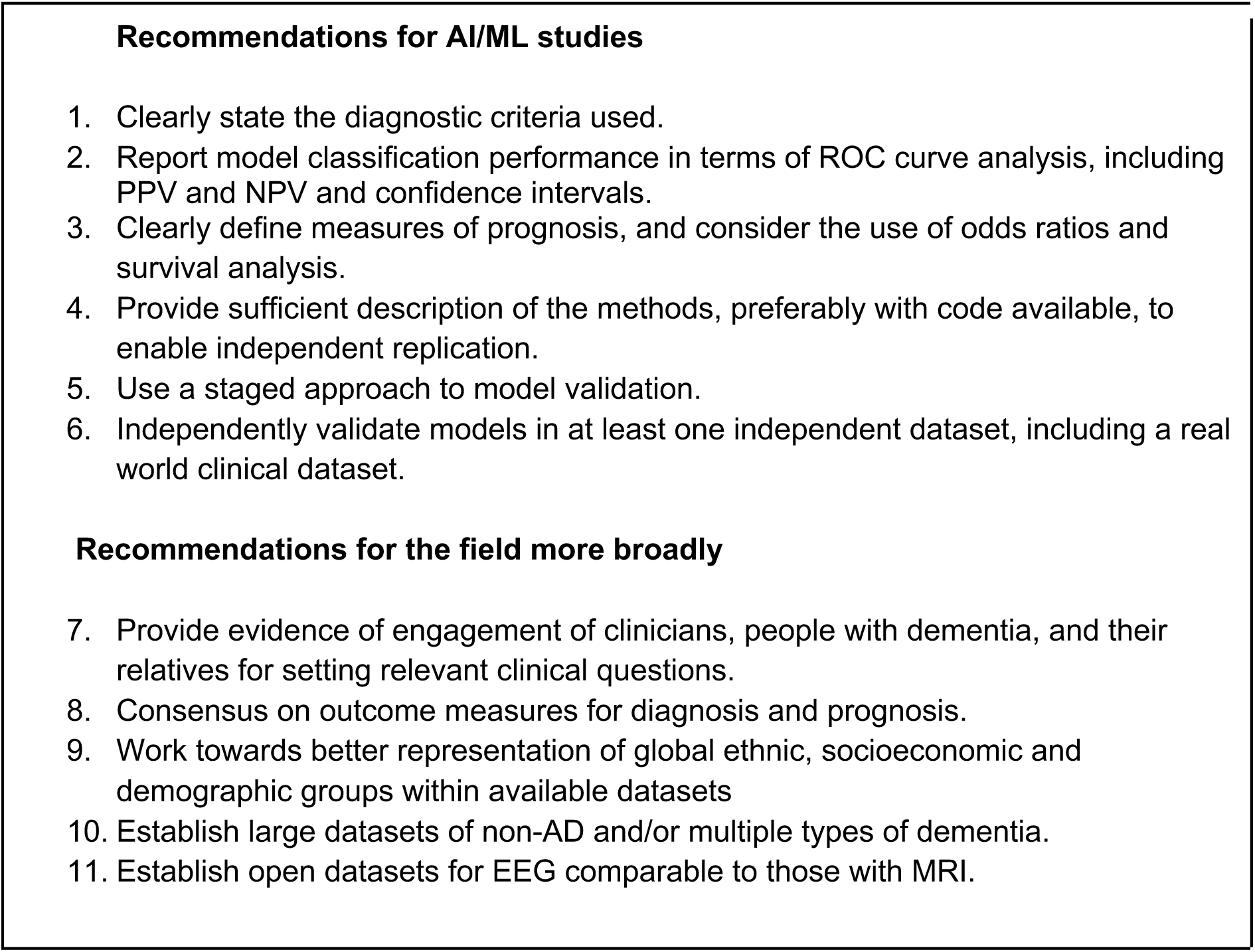
Recommendations to move towards clinically useful, AI/ML methods applied to neuroimaging for dementia.

|  |
| --- |
| **Recommendations for AI/ML studies**<br>1. Clearly state the diagnostic criteria used.<br>2. Report model classification performance in terms of ROC curve analysis, including PPV and NPV and confidence intervals.<br>3. Clearly define measures of prognosis, and consider the use of odds ratios and survival analysis.<br>4. Provide sufficient description of the methods, preferably with code available, to enable independent replication.<br>5. Use a staged approach to model validation.<br>6. Independently validate models in at least one independent dataset, including a real world clinical dataset.<br>**Recommendations for the field more broadly**<br>7. Provide evidence of engagement of clinicians, people with dementia, and their relatives for setting relevant clinical questions.<br>8. Consensus on outcome measures for diagnosis and prognosis.<br>9. Work towards better representation of global ethnic, socioeconomic and demographic groups within available datasets<br>10. Establish large datasets of non-AD and/or multiple types of dementia.<br>11. Establish open datasets for EEG comparable to those with MRI. |

We anticipate that the implementation of these recommendations for future studies using AI algorithms for diagnosis and prognosis in dementia will help move the field towards a future where these AI/ML tools can be used by clinicians to benefit patients. Even with these recommendations, several practical considerations remain including integration into clinical workflows and overcoming regulatory and financial barriers.

### Limitations

This systematic review has three main limitations. First, although we aimed to provide an informed and broad overview of the existing literature on this subject, our exclusion of reports not written in English and those where the full-text was not available meant that some reports which would have otherwise met the inclusion criteria may not have been covered in this review. Two key additional exclusion criteria were the decisions not to include studies using linear regression for classification, and studies combining neuroimaging with other biomarkers. Our motivation was to focus specifically on neuroimaging, and specifically on recognised AI/ML methods, but it is possible we excluded papers with high clinical value and translational potential.

Second, the heterogeneity in classification tasks, ML methods used and statistical reporting across reports may have introduced bias when trying to decipher which tasks and results to extract. More specifically, this was an issue with the more technical reports which compared multiple (often >5) machine learning methods across three or more classification groups introducing a large number of comparisons and results to consolidate and extract. We do not address significant ethical issues in big data analysis of data security, consent to data sharing, and the acceptability of AI methods to clinicians and the general public.

Third, we employed a risk of bias screening tool that depended on a subjective judgment for each paper’s inclusion or exclusion. We chose a low threshold for inclusion based on study quality in order to accurately depict and identify current barriers in the literature limiting translation to clinical practice. We only excluded studies exhibiting clear methodological concerns, such as lack of reporting of basic participant demographics.

### Conclusions

In this structured review, we generate a number of recommendations to facilitate translation of AI/ML methods for patient benefit in the diagnosis and prognosis of dementia. We highlight the need for early clinical involvement, methodological heterogeneity, and appropriate and representative data as three key areas to address for the field to move towards translation. We believe that addressing these and additional logistical and regulatory issues will improve patient care. It will involve collaboration between clinical academics and machine learning experts, validation of AI tools through open source data sharing and certification as well as large-scale initiatives to collate representative datasets.

## Individual funding

Jose Bernal is supported by the MRC Doctoral Training Programme in Precision Medicine (Award Reference No. 2096671).

Amanpreet Badhwar is supported by Fonds de recherche du Québec Santé - Chercheur boursiers Junior 1 and Fondation Courtois.

Matthew Betts is supported by the Deutsche Forschungsgemeinschaft (DFG, German Research Foundation) – Project-ID 425899996 – SFB 1436 Project A08 and by the German Federal Ministry of Education and Research (BMBF, funding code 01ED2102B) under the aegis of JPND.

Eugene Tang, NIHR Clinical Lecturer, is funded by the National Institute for Health Research (NIHR). The views expressed in this publication are those of the author(s) and not necessarily those of the NIHR, NHS or the UK Department of Health and Social Care

Sofia Michopoulou, NIHR Clinical Lecturer, was funded by the National Institute for Health Research (NIHR), the NIHR Applied Research Collaboration ARC Wessex, the Southampton Academy of Research and the Health Education England Topol Fellowship program. The views expressed in this publication are those of the author and not necessarily those of the funding bodies.

Carlos Muñoz-Neira was supported by the Government of Chile through ’Becas Chile’ and CONICYT - National Commission for Scientific and Technological Research [CONICYT - Comisión Nacional de Investigación Científica y Tecnológica], the University of Bristol (Grant Code G100030-150), and its Postdoctoral Research Associate position at the University of Sheffield.

## Data Availability

Data produced in the present work are contained in the manuscript and supplementary material.

