## Supplementary material 1 for "Artificial intelligence for diagnosis and prognosis in neuroimaging for dementia; a systematic review"

1. Exact search terms used in each database:

**Medline via Ovid**

Dementia.ti,ab,kw. or exp Dementia/

AND

(Alzheimer* or “Lewy bod*” or “frontotemporal lobar degenerat*” or “frontotemporal dementia” or “progressive supranuclear palsy” or Huntington* or “corticobasal degeneration”).ti,ab,kw. or exp alzheimer disease/ or exp dementia, vascular/

AND

((cogni* adj3 (declin* or function* or dysfunction*)) or (mental adj3 (declin* or function* or dysfunction*))).ti,ab,kw. or (memor* or psychometric* or neuropsycholog*).ti,ab,kw. or exp Memory/ or exp Neuropsychological Tests/ or Cognitive Dysfunction/ or exp mild cognitive impairment/

AND

(“Artificial Intelligence” OR AI OR “neural network*” OR “deep learning” OR “machine learning” OR ML OR “ridge regression” OR “least absolute shrinkage and selection operator” OR LASSO OR “elastic net” OR classifier OR “support vector machine” OR “random forest*” OR “nearest-neighbo*” OR markov OR clustering OR convolution OR segmentation OR prediction OR regression OR classification OR “decision support system*” OR automl OR ensemble OR unsupervised OR supervised OR “dimensionality reduction”).ti,ab,kw. OR Artificial Intelligence/ or Machine Learning/ or Deep Learning/ or Neural networks, computer/ OR support vector machine/ or Markov chains/ or regression analysis/ or cluster analysis/ or regression analysis/

AND

(Neuroimaging OR “Magnetic Resonance Imaging” OR MRI OR “Positron Emission Tomography” OR PET OR Ultrasound OR “structural imaging” OR “functional imaging” OR fMRI OR “dynamic imaging” OR “DCE-MRI” OR “Single Photon Emission Computed Tomography” OR SPECT OR Electroencephalography OR EEG OR Magnetoencephalography OR MEG).ti,ab,kw. OR neuroimaging/ or magnetic resonance imaging/ or positron-emission tomography/ or Tomography, Emission-Computed, Single-Photon/ or Magnetoencephalography/ or Electroencaphalography/

AND

(Diagnosis OR Prognosis).ti,ab,kw. or diagnosis/ or prognosis/

**Embase (via Ovid)**

Dementia.ti,ab,kw. or exp Dementia/

AND

(Alzheimer* or “Lewy bod*” or “frontotemporal lobar degenerat*” or “frontotemporal dementia” or “progressive supranuclear palsy” or Huntington* or “corticobasal degeneration").ti,ab,kw. or exp alzheimer disease/ or exp dementia, vascular/ or huntinton chorea/ or corticobasal degeneration/

AND

((cogni* adj3 (declin* or function* or dysfunction*)) or (mental adj3 (declin* or function* or dysfunction*))).ti,ab,kw. or (memor* or psychometric* or neuropsycholog*).ti,ab,kw. or exp Memory/ or exp Neuropsychological Test/ or Cognitive Defect/ or exp mild cognitive impairment/

AND

("Artificial Intelligence" OR AI OR "neural network*" OR "deep learning" OR "machine learning" OR ML OR "ridge regression" OR "least absolute shrinkage and selection operator" OR LASSO OR "elastic net" OR classifier OR "support vector machine" OR "random forest*" OR "nearest-neighbo*" OR markov OR clustering OR convolution OR segmentation OR prediction OR regression OR classification OR "decision support system*" OR automl OR ensemble OR unsupervised OR supervised OR "dimensionality reduction").ti,ab,kw. or artificial intelligence/ or artificial neural network/ or deep learning/ or machine learning/ or classifier/ or support vector machine/ or random forest/ or k nearest neighbor/ or Markov chain/ or clustering algorithm/ or convolution algorithm/ or segmentation algorithm/ or regression analysis/ or dimensionality reduction/

AND

(Neuroimaging OR "Magnetic Resonance Imaging" OR MRI OR "Positron Emission Tomography" OR PET OR Ultrasound OR "structural imaging" OR "functional imaging" OR fMRI OR "dynamic imaging" OR "DCE-MRI" OR "Single Photon Emission Computed Tomography" OR SPECT OR Electroencephalography OR EEG OR Magnetoencephalography OR MEG).ti,ab,kw. or neuroimaging/ or nuclear magnetic resonance imaging/ or positron emission tomography/ or ultrasound/ or Single Photon Emission Computed Tomography/ or Electroencaphalography/ or Magnetoencephalography/

AND

(Diagnosis OR Prognosis).ti,ab,kw. or diagnosis/ or prognosis/

**Cochrane Library**

Dementia:ti,ab,kw or MeSH descriptor: [Dementia] explode all trees

AND

(Alzheimer* or "Lewy bod*" or "frontotemporal lobar degenerat*" or "frontotemporal dementia" or "progressive supranuclear palsy" or Huntington* or "corticobasal degeneration"):ti,ab,kw or MeSH descriptor: [alzheimer disease] explode all trees or MeSH descriptor: [dementia, vascular] explode all trees or MeSH descriptor: [Huntington disease] explode all trees

AND

((cogni* Near/3 (declin* or function* or dysfunction*)) or (mental Near/3 (declin* or function* or dysfunction*))):ti,ab,kw or (memor* or psychometric* or neuropsycholog*):ti,ab,kw or MeSH descriptor: [Memory] explode all trees or MeSH descriptor: [Neuropsychological Tests] this term only or MeSH descriptor: [Cognitive Dysfunction] this term only

AND

(“Artificial Intelligence” OR AI OR “neural network*” OR “deep learning” OR “machine learning” OR ML OR “ridge regression” OR “least absolute shrinkage and selection operator” OR LASSO OR “elastic net” OR classifier OR “support vector machine” OR “random forest*” OR “nearest-neighbo*” OR markov OR clustering OR convolution OR segmentation OR prediction OR regression OR classification OR “decision support system*” OR automl OR ensemble OR unsupervised OR supervised OR “dimensionality reduction”):ti,ab,kw or MeSH descriptor: [Artificial intelligence] this term only or MeSH descriptor: [neural networks, computer] this term only or MeSH descriptor: [deep learning] this term only or MeSH descriptor: [machine learning] this term only or MeSH descriptor: [support vector machine] or MeSH descriptor: [markov chains] this term only or MeSH descriptor: [cluster analysis] this term only or MeSH descriptor: [regression analysis] this term only or MeSH descriptor: [Multifactor dimensionality reduction] this term only

AND

(Neuroimaging OR “Magnetic Resonance Imaging” OR MRI OR “Positron Emission Tomography” OR PET OR Ultrasound OR “structural imaging” OR “functional imaging” OR fMRI OR “dynamic imaging” OR “DCE-MRI” OR “Single Photon Emission Computed Tomography” OR SPECT OR Electroencephalography OR EEG OR Magnetoencephalography OR MEG):ti,ab,kw or MeSH descriptor: [neuroimaging] this term only or MeSH descriptor: [functional neuroimaging] this term only or MeSH descriptor: [positron-emission tomography] this term only or MeSH descriptor: [ultrasonography] this term only or MeSH descriptor: [Tomography, Emission-Computed, Single-Photon] this term only or MeSH descriptor: [Magnetoencephalography] this term only

AND

(Diagnosis OR Prognosis):ti,ab,kw or MeSH descriptor: [diagnosis] this term only or MeSH descriptor: [prognosis] this term only

**BNI (via ProQuest)**

ti(Dementia) or ab(Dementia) or MAINSUBJECT.EXACT("Dementia")

AND

ti(Alzheimer* or “Lewy bod*” or “frontotemporal lobar degenerat*” or “frontotemporal dementia” or “progressive supranuclear palsy” or Huntington* or “corticobasal degeneration”) or ab(Alzheimer* or “Lewy bod*” or “frontotemporal lobar degenerat*” or “frontotemporal dementia” or “progressive supranuclear palsy” or Huntington* or “corticobasal degeneration”) or MAINSUBJECT.EXACT("Alzheimers disease") or MAINSUBJECT.EXACT("Huntingtons disease")

AND

ti((cogni* Near/3 (declin* or function* or dysfunction*)) or (mental Near/3 (declin* or function* or dysfunction*))) or ti(memor* or psychometric* or neuropsycholog*) or ab((cogni* Near/3 (declin* or function* or dysfunction*)) or (mental Near/3 (declin* or function* or dysfunction*))) or ab(memor* or psychometric* or neuropsycholog*) or AB (memor* or psychometric* or neuropsycholog*) or MAINSUBJECT.EXACT("Memory")

AND

ti(“Artificial Intelligence” OR AI OR “neural network*” OR “deep learning” OR “machine learning” OR ML OR “ridge regression” OR “least absolute shrinkage and selection operator” OR LASSO OR “elastic net” OR classifier OR “support vector machine” OR “random forest*” OR “nearest-neighbo*” OR markov OR clustering OR convolution OR segmentation OR prediction OR regression OR classification OR “decision support system*” OR automl OR ensemble OR unsupervised OR supervised OR “dimensionality reduction”) OR ab(“Artificial Intelligence” OR AI OR “neural network*” OR “deep learning” OR “machine learning” OR ML OR “ridge regression” OR “least absolute shrinkage and selection operator” OR LASSO OR “elastic net” OR classifier OR “support vector machine” OR “random forest*” OR “nearest-neighbo*” OR markov OR clustering OR convolution OR segmentation OR prediction OR regression OR classification OR “decision support system*” OR automl OR ensemble OR unsupervised OR supervised OR “dimensionality reduction”) or MAINSUBJECT.EXACT("Artificial intelligence") or MAINSUBJECT.EXACT("Neural networks") or MAINSUBJECT.EXACT("Deep learning") or MAINSUBJECT.EXACT("Machine learning") or MAINSUBJECT.EXACT("Support vector machines") or MAINSUBJECT.EXACT("Clustering") or MAINSUBJECT.EXACT("Regression analysis") or MAINSUBJECT.EXACT("Decision support systems")

AND

ti(Neuroimaging OR “Magnetic Resonance Imaging” OR MRI OR “Positron Emission Tomography” OR PET OR Ultrasound OR “structural imaging” OR “functional imaging” OR fMRI OR “dynamic imaging” OR “DCE-MRI” OR “Single Photon Emission Computed Tomography” OR SPECT OR Electroencephalography OR EEG OR Magnetoencephalography OR MEG) or ab(Neuroimaging OR “Magnetic Resonance Imaging” OR MRI OR “Positron Emission Tomography” OR PET OR Ultrasound OR “structural imaging” OR “functional imaging” OR fMRI OR “dynamic imaging” OR “DCE-MRI” OR “Single Photon Emission Computed Tomography” OR SPECT OR Electroencephalography OR EEG OR Magnetoencephalography OR MEG) OR MAINSUBJECT.EXACT("Medical imaging") or MAINSUBJECT.EXACT("Medical imaging") or MAINSUBJECT.EXACT("Ultrasonic imaging") or MAINSUBJECT.EXACT("Electroencephalography")

AND

ti(Diagnosis OR Prognosis) or ab(Diagnosis OR Prognosis) or MAINSUBJECT.EXACT("Medical prognosis") or MAINSUBJECT.EXACT("Medical diagnosis")

**PsycInfo (via Ebscohost)**

TI (Dementia) or AB (Dementia) or KW (Dementia) or DE “Dementia”

AND

TI (Alzheimer* or “Lewy bod*” or “frontotemporal lobar degenerat*” or “frontotemporal dementia” or “progressive supranuclear palsy” or Huntington*” or “corticobasal degeneration”) or AB (Alzheimer* or “Lewy bod*” or “frontotemporal lobar degenerat*” or “frontotemporal dementia” or “progressive supranuclear palsy” or Huntington* or “corticobasal degeneration”) or KW (Alzheimer* or “Lewy bod*” or “frontotemporal lobar degenerat*” or “frontotemporal dementia” or “progressive supranuclear palsy” or Huntington* or “corticobasal degeneration”) or DE “Alzheimer’s Disease” or DE “dementia with lewy bodies” or DE “Huntingtons Disease” or DE “corticobasal degeneration”

AND

TI ((cogni* N3 (declin* or function* or dysfunction*)) or (mental N3 (declin* or function* or dysfunction*))) or TI (memor* or psychometric* or neuropsycholog*) or AB ((cogni* N3 (declin* or function* or dysfunction*)) or (mental N3 (declin* or function* or dysfunction*))) or AB (memor* or psychometric* or neuropsycholog*) or AB (memor* or psychometric* or neuropsycholog*) or KW ((cogni* N3 (declin* or function* or dysfunction*)) or (mental N3 (declin* or function* or dysfunction*))) or KW (memor* or psychometric* or neuropsycholog*) or DE “mild cognitive impairment” or DE “memory” or DE “cognitive impairment”

AND

TI (“Artificial Intelligence” OR AI OR “neural network*” OR “deep learning” OR “machine learning” OR ML OR “ridge regression” OR “least absolute shrinkage and selection operator” OR LASSO OR “elastic net” OR classifier OR “support vector machine” OR “random forest*” OR “nearest-neighbo*” OR markov OR clustering OR convolution OR segmentation OR prediction OR regression OR classification OR “decision support system*” OR automl OR ensemble OR unsupervised OR supervised OR “dimensionality reduction”) OR AB (“Artificial Intelligence” OR AI OR “neural network*” OR “deep learning” OR “machine learning” OR ML OR “ridge regression” OR “least absolute shrinkage and selection operator” OR LASSO OR “elastic net” OR classifier OR “support vector machine” OR “random forest*” OR “nearest-neighbo*” OR markov OR clustering OR convolution OR segmentation OR prediction OR regression OR classification OR “decision support system*” OR automl OR ensemble OR unsupervised OR supervised OR “dimensionality reduction”) or KW (“Artificial Intelligence” OR AI OR “neural network*” OR “deep learning” OR “machine learning” OR ML OR “ridge regression” OR “least absolute shrinkage and selection operator” OR LASSO OR “elastic net” OR classifier OR “support vector machine” OR “random forest*” OR “nearest-neighbo*” OR markov OR clustering OR convolution OR segmentation OR prediction OR regression OR classification OR “decision support system*” OR automl OR ensemble OR unsupervised OR supervised OR “dimensionality reduction”) or DE “artificial intelligence” or DE “neural networks” or DE “machine learning” or DE “markov chains” or DE “cluster analysis” or DE “decision support systems”

AND

TI (Neuroimaging OR “Magnetic Resonance Imaging” OR MRI OR “Positron Emission Tomography” OR PET OR Ultrasound OR “structural imaging” OR “functional imaging” OR fMRI OR “dynamic imaging” OR “DCE-MRI” OR “Single Photon Emission Computed Tomography” OR SPECT OR Electroencephalography OR EEG OR Magnetoencephalography OR MEG) or AB (Neuroimaging OR “Magnetic Resonance Imaging” OR MRI OR “Positron Emission Tomography” OR PET OR Ultrasound OR “structural imaging” OR “functional imaging” OR fMRI OR “dynamic imaging” OR “DCE-MRI” OR “Single Photon Emission Computed Tomography” OR SPECT OR Electroencephalography OR EEG OR Magnetoencephalography OR MEG) or KW (Neuroimaging OR “Magnetic Resonance Imaging” OR MRI OR “Positron Emission Tomography” OR PET OR Ultrasound OR “structural imaging” OR “functional imaging” OR fMRI OR “dynamic imaging” OR “DCE-MRI” OR “Single Photon Emission Computed Tomography” OR SPECT OR Electroencephalography OR EEG OR Magnetoencephalography OR MEG) or DE “Neuroimaging” or DE “magnetic resonance imaging” or DE “positron emission tomography” or DE “ultrasound” or DE “functional magnetic resonance imaging” or DE "Electroencephalography" or DE “Magnetoencephalography”

AND

TI (Diagnosis OR Prognosis) or AB (Diagnosis OR Prognosis) or KW (Diagnosis OR Prognosis) or DE “diagnosis” or DE “prognosis”

**Cinahl (via Ebscohost)**

TI (Dementia) or AB (Dementia) or (MH “dementia+”)

AND

TI (Alzheimer* or “Lewy bod*” or “frontotemporal lobar degenerat*” or “frontotemporal dementia” or “progressive supranuclear palsy” or Huntington* or “corticobasal degeneration”) or AB (Alzheimer* or “Lewy bod*” or “frontotemporal lobar degenerat*” or “frontotemporal dementia” or “progressive supranuclear palsy” or Huntington* or “corticobasal degeneration”) or (MH “Alzheimer’s Disease+”) or (MH “Lewy Body Disease”) or (MH “Huntington’s Disease”)

AND

TI ((cogni* N3 (declin* or function* or dysfunction*)) or (mental N3 (declin* or function* or dysfunction*))) or TI (memor* or psychometric* or neuropsycholog*) or AB ((cogni* N3 (declin* or function* or dysfunction*)) or (mental N3 (declin* or function* or dysfunction*))) or AB (memor* or psychometric* or neuropsycholog*) or (MH “memory+”)

AND

TI (“Artificial Intelligence” OR AI OR “neural network*” OR “deep learning” OR “machine learning” OR ML OR “ridge regression” OR “least absolute shrinkage and selection operator” OR LASSO OR “elastic net” OR classifier OR “support vector machine” OR “random forest*” OR “nearest-neighbo*” OR markov OR clustering OR convolution OR segmentation OR prediction OR regression OR classification OR “decision support system*” OR automl OR ensemble OR unsupervised OR supervised OR “dimensionality reduction”) OR AB (“Artificial Intelligence” OR AI OR “neural network*” OR “deep learning” OR “machine learning” OR ML OR “ridge regression” OR “least absolute shrinkage and selection operator” OR LASSO OR “elastic net” OR classifier OR “support vector machine” OR “random forest*” OR “nearest-neighbo*” OR markov OR clustering OR convolution OR segmentation OR prediction OR regression OR classification OR “decision support system*” OR automl OR ensemble OR unsupervised OR supervised OR “dimensionality reduction”) or (MH “Artificial Intelligence”) OR (MH “neural networks (computer)”) or (MH “deep learning”) or (MH “machine learning”) or (MH “support vector machine”) or (MH “random forest”) or (MH “cluster analysis”)

AND

TI (Neuroimaging OR “Magnetic Resonance Imaging” OR MRI OR “Positron Emission Tomography” OR PET OR Ultrasound OR “structural imaging” OR “functional imaging” OR fMRI OR “dynamic imaging” OR “DCE-MRI” OR “Single Photon Emission Computed Tomography” OR SPECT OR Electroencephalography OR EEG OR Magnetoencephalography OR MEG) or AB (Neuroimaging OR “Magnetic Resonance Imaging” OR MRI OR “Positron Emission Tomography” OR PET OR Ultrasound OR “structural imaging” OR “functional imaging” OR fMRI OR “dynamic imaging” OR “DCE-MRI” OR “Single Photon Emission Computed Tomography” OR SPECT OR Electroencaphalograph OR EEG OR Magnetoencephalography OR MEG) or (MH “neuroradiography”) or (MH “Magnetic Resonance Imaging”) or (MH “tomography, emission-computed”) or (MH “ultrasonography”) or (MH "Tomography, Emission-Computed, Single-Photon") or (MH “Electroencephalography”)

AND

TI (Diagnosis OR Prognosis) or AB (Diagnosis OR Prognosis) OR (MH “diagnosis”) or (MH “prognosis”)

**Emcare (via Ovid)**

Dementia.ti,ab,kw. or exp Dementia/

AND

(Alzheimer* or “Lewy bod*” or “frontotemporal lobar degenerat*” or “frontotemporal dementia” or “progressive supranuclear palsy” or Huntington* or “corticobasal degeneration”).ti,ab,kw. or diffuse Lewy body disease/ or exp alzheimer disease/ or multiinfarct dementia/ or huntington chorea/ or corticobasal degeneration/ or progressive supranuclear palsy/

AND

((cogni* adj3 (declin* or function* or dysfunction*)) or (mental adj3 (declin* or function* or dysfunction*))).ti,ab,kw. or (memor* or psychometric* or neuropsycholog*).ti,ab,kw. or exp Memory/ or exp Neuropsychological Tests/ or Cognitive Defect/ or or exp mild cognitive impairment/

AND

(“Artificial Intelligence” OR AI OR “neural network*” OR “deep learning” OR “machine learning” OR ML OR “ridge regression” OR “least absolute shrinkage and selection operator” OR LASSO OR “elastic net” OR classifier OR “support vector machine” OR “random forest*” OR “nearest-neighbo*” OR markov OR clustering OR convolution OR segmentation OR prediction OR regression OR classification OR “decision support system*” OR automl OR ensemble OR unsupervised OR supervised OR “dimensionality reduction”).ti,ab,kw. or Artificial Intelligence/ or artificial neural network/ or deep learning/ or machine learning/ or classifier/ or support vector machine/ or random forest/ or k nearest-neighbor/ or markov chain/ or cluster analysis/ or convolution algorithm/ or segmentation algorithm/ or regression analysis/ or classification algorithm/ or decision support system/ or dimensionality reduction/

AND

(Neuroimaging OR “Magnetic Resonance Imaging” OR MRI OR “Positron Emission Tomography” OR PET OR Ultrasound OR “structural imaging” OR “functional imaging” OR fMRI OR “dynamic imaging” OR “DCE-MRI” OR “Single Photon Emission Computed Tomography” OR SPECT OR Electroencephalography OR EEG OR Magnetoencephalography OR MEG).ti,ab,kw. or neuroimaging/ or functional neuroimaging/ or nuclear magnetic resonance imaging/ or Positron Emission Tomography/ or ultrasound/ or Single Photon Emission Computed Tomography/ or Electroencephalography/ or Magnetoencephalography/

AND

(Diagnosis OR Prognosis).ti,ab,kw. or diagnosis/ or prognosis/

2. Spreadsheet with final 252 reports included (separate spreadsheet)

3. Methods and weights used for the Stratified Weighted average Sum (SWS) approach:

We constructed a Stratified Weighted average Sum (SWS) by assigning weights at (a) Datasets, and (b) Imaging modality levels which is outlined below.

**Working principle of Stratified Weighted average Sum** (**SWS)**: It is defined as SWS := (Dataset_weight)*(Img_modality_weight)*(Avg_AUC); where (i) Dataset_weight is number of studies used a specific type of datasets/Total number of studies (table(a)), (ii) Img_modality_weight is number of studies used subtypes within each dataset type/Total number of studies (table(b)), and (iii) Avg_AUC is average of AUC estimates found after doing stratification of imaging modality data.

The calculated variance of AUC estimates on a specific type of datasets is lower compared to variance of AUC estimations on unsegregated datasets. The variance decreases further when calculated on imaging modality (subtypes within each dataset type). Which means that SWS not only reduces the heterogeneity factor involved in the AUC estimation but also weights the AUC estimates based on the number of studies used which type of dataset and modality.

Note: If SWS matrix is not sparse then SWS AUC value will be close to Avg_AUC otherwise less/very less than Avg_AUC for a sparse/very sparse SWS matrix. Moreover the sparsity also tells us that the studies were mostly focused on which type/subtype of data and which ones were ignored completely.

Table(a)

| Dataset | n | Weight |
|---|---|---|
| Only ADNI | 163 | 0.649402 |
| Local data | 57 | 0.227092 |
| ADNI + Others | 15 | 0.059761 |
| Only OASIS | 5 | 0.0199203 |

| | | |
|---|---|---|
| Only BLSA | 2 | 0.00796813 |
| Only Bdx-3C | 1 | 0.00398406 |
| Other | 8 | 0.0318725 |

Table(b)

| Imaging Modality | n | Weight |
|---|---|---|
| Only sMRI | 132 | 0.52381 |
| Only fMRI | 19 | 0.0753968 |
| Only PET | 27 | 0.107143 |
| Only EEG | 22 | 0.0873016 |
| Only DTI | 1 | 0.00396825 |
| Only SPECT | 2 | 0.00793651 |
| Others | 49 | 0.194444 |

Results:
The following tables (R.a), (R.b), and (R.c) depict the total sums of the respective SWS analyses including the algorithmic (SWS=0.585457), discriminative (SWS=0.601972), and generative (SWS=0.555611) models.

(R.a) **Algorithmic models**:

| Data_Source | Weighted AUC | | | | | | |
|---|---|---|---|---|---|---|---|
| | sMRI | fMRI | PET | EEG | DTI | SPECT | Others |
| Only ADNI | 0.318465 | 0.043969 | 0.065216 | 0 | | 0 | 0 |
| Local data | 0.115979 | 0 | 0.021533 | 0.004017 | | 0 | 0 |
| ADNI+Others | 0 | 0 | 0 | 0 | | 0 | 0 |
| OASIS | 0 | 0 | 0 | 0 | | 0 | 0 |
| BLSA | 0 | 0 | 0 | 0 | | 0 | 0 |
| Bdx-3C | 0 | 0 | 0 | 0 | | 0 | 0 |

| | | | | | | | |
|---|---|---|---|---|---|---|---|
| Others | 0.016278 | 0 | 0 | 0 | | 0 | 0 |

(R.b) **Discriminative models**:

| Data_Source | Weighted AUC | | | | | | |
|---|---|---|---|---|---|---|---|
| | sMRI | fMRI | PET | EEG | DTI | SPECT | Others |
| Only ADNI | 0.321962 | 0.046515 | 0.066767 | 0 | 0.002 422 | 0 | 0 |
| Local data | 0.113481 | 0.017122 | 0 | 0 | 0 | 0 | 0 |
| ADNI+Others | 0.024104 | 0 | 0 | 0 | 0 | 0 | 0 |
| OASIS | 0.009600 | 0 | 0 | 0 | 0 | 0 | 0 |
| BLSA | 0 | 0 | 0 | 0 | 0 | 0 | 0 |
| Bdx-3C | 0 | 0 | 0 | 0 | 0 | 0 | 0 |
| Others | 0 | 0 | 0 | 0 | 0 | 0 | 0 |

(R.c) **Generative models**:
.

| Data_Source | Weighted AUC | | | | | | |
|---|---|---|---|---|---|---|---|
| | sMRI | fMRI | PET | EEG | DTI | SPECT | Others |
| Only ADNI | 0.312091 | 0.039170 | 0.060534 | 0 | | 0 | 0 |
| Local data | 0.109793 | 0.015804 | 0 | 0 | | 0 | 0 |
| ADNI+Others | 0 | 0 | 0 | 0 | | 0 | 0 |
| OASIS | 0 | 0 | 0 | 0 | | 0 | 0 |
| BLSA | 0 | 0 | 0 | 0 | | 0 | 0 |
| Bdx-3C | 0.001523 | 0 | 0 | 0 | | 0 | 0 |
| Others | 0.016695 | 0 | 0 | 0 | | 0 | 0 |
